# Variation in the Incidence of ventriculostomy related infection in critically ill patients

**DOI:** 10.1101/2022.04.19.22273488

**Authors:** Sara N. Bassin, David H. Tian, Simon Chadwick, Sajeev Mahendran, Oliver Flower, Emily Fitzgerald, Jonathon Parkinson, Archie Darbar, Pierre Janin, Anthony Delaney

## Abstract

**Introduction:** Ventriculostomy related infection (VRI) or ventriculitis is a common and serious complication related to the placement of an external ventricular drain. Numerous sets of diagnostic criteria for VRI have been reported. We sought to estimate the variation in the incidence of VRI in a cohort of patients according to published diagnostic criteria.

**Materials and Methods:** We conducted a retrospective cohort study. We included adult patients admitted to the Neuroscience intensive care unit with traumatic brain injury (TBI), subarachnoid haemorrhage (SAH) and intracerebral haemorrhage (ICH) who required an EVD. We estimated the incidence of VRI according to published diagnostic criteria. We compared the incidence to clinicians’ diagnoses of VRI. The primary outcome was the estimated incidence of VRI.

**Results:** There were 190 study participants, median age (interquartile range) of 58 (48 - 72), 106 (55.8%) were female. Admitting diagnoses was ICH in 30 (15.8%), TBI in 49 (25.8%) and SAH in 111 (58.4%) of cases respectively. There were 158 (83.2%) who required mechanical ventilation for a median of 6 (2-13) days. There were 29 (15.3%) who were treated for VRI by clinicians, with 6 (3.2%) having a positive culture. Variation in the diagnostic criteria led to an estimated incidence of VRI that ranged from 1 (0.5%) to 178 (93.7%).

**Conclusion:** In this critically ill cohort, the estimated incidence of VRI varied widely depending on which diagnostic criteria for VRI were applied. A comprehensive, consistent, objective and universal set of diagnostic criteria for ventriculostomy related infection is needed.

## Introduction

The insertion of an external ventricular drain (EVD) is one of the most common neurosurgical procedures, generally undertaken for the treatment of acute hydrocephalus or for monitoring intracranial pressure.^1^ While the insertion of an EVD may be life-saving, the procedure has known complications including infection, haemorrhage and misplacement of the catheter.^2^ Infection, referred to as ventriculostomy related infection (VRI) is the most common and clinically significant complication of EVD placement.^3^ Patients with VRI appear to have an increased risk of morbidity,^4^ mortality,^5^ increased Intensive Care Unit (ICU) and hospital length of stay, with increased cost of care.^6^

Strategies to ameliorate the adverse consequences of VRI are required and guidelines for the management of EVDs have been produced.^7^ A key difficulty in providing robust evidence based-recommendations for the prevention of VRI is that there is no consistent set of diagnostic criteria for VRI.^7^ A systematic review in 2016 reported identifying 17 separate published definitions of VRI, with an estimate of the incidence of VRI in a test cohort that ranged from 22% to 94%.^8^ An underreported issue confronting clinicians attempting to diagnose VRI in in a critically ill population is that many definitions of VRI require clinical parameters such as the presence of headache, neck stiffness or altered level of consciousness that are unreliable due to the confounding effects of the primary condition that necessitated the insertion of the EVD (e.g., subarachnoid haemorrhage) or by the treatments provided to patients with acute severe brain injury, such as tracheal intubation and the provision of intravenous sedative and analgesic agents.^9,10^

The lack of clarity in the diagnostic criteria for VRI creates problems for clinicians. While underdiagnosis may lead to missed opportunities to intervene and reduce preventable morbidity and mortality,^11^ over treatment is associated with potential adverse events.^12^ It is not clear how the various diagnostic criteria for VRI are implemented in routine clinical practice. Therefore, we conducted a retrospective observational study to assess the variation between the incidence of VRI diagnosed and treated in the clinical setting and the estimated incidence of VRI using the published diagnostic criteria.^8^

## Materials and methods

### Study design and setting

This retrospective cohort study was conducted in the Neuroscience Intensive Care Unit at the Royal North Shore Hospital. The Royal North Shore Hospital is a 600-bed referral centre in Sydney, Australia. The Neuroscience Intensive Care Unit is a dedicated 13 bed Neurocritical care unit that admits approximately 800 patients per year. Clinical and laboratory data from patients with traumatic brain injury (TBI), subarachnoid haemorrhage (SAH) and intracerebral haemorrhage (ICH) admitted to the Neuroscience ICU are collected in a prospective registry, the Neurological Outcomes in Intensive Care (NOICE) registry, which was used for the conduct of this study. ^13^ Data regarding CSF microbiology and biochemistry were extracted separately. Ethics approval to conduct the study was obtained from the Northern Sydney Local Health District Human Research Ethics Committee (RESP/18/339). The report was structured according to the guidelines suggested by the STROBE statement.^14^

### Participants

We included data from adult patients admitted to the Neuroscience ICU from January 1^st^ 2015 to July 22^nd^ 2018 with a TBI, SAH or ICH, who had an EVD placed for a clinical indication and had at least one CSF specimen collected.

### Data Sources and Variables

Data collected from the NOICE registry included patient demographics and comorbidities, dates of hospital, ICU admission and discharge, primary neurosurgical pathology, illness severity scoring, clinical and laboratory data on admission and daily measures, treatment modalities, and complications of ICU stay. We also collected data to grade the severity of illness for each condition. For patients with SAH, three scoring systems were used; The World Federation of Neurosurgical Societies clinical grading system,^15^ and the Fisher and Claassen radiological grading systems.^16^ For patients with TBI, the Marshall CT classification system was used.^17^ In the patients who had an ICH, the ICH score was used.^18^ We collected data regarding the haematological and biochemical profile of any CSF specimens as well as the results of microscopy and culture results.

A confirmed clinical diagnosis of VRI was recorded in our cohort for patients when there was growth of a pathogenic organism in the CSF cultures. A treated clinical case of VRI was recorded when a patient was recorded as receiving intravenous antibiotics due to clinical diagnosis of VRI. A suspected case of VRI was recorded when the CSF white cell count: red cell count ratio was >1:200.

The estimated incidence of VRI according to our clinical criteria was compared to the estimated incidence of VRI obtained by using the various criteria set out in the published literature.^8^ The full details of the 17 sets of diagnostic criteria for ventriculitis can be found in Appendix 1. From each set of diagnostic criteria, we used the published thresholds for CSF parameters including white cell count (WCC), red cell count (RCC) to WCC ratio, CSF protein and glucose concentration, physiological variables (including temperature), and laboratory values such as peripheral WCC. When parameter cut-offs were not specified we used the abnormal CSF and laboratory parameters outlined in the Royal College of Pathologists of Australasia manual (CSF cell count < 5× 10^6^/L mononuclears; no neutrophils or red cells, glucose 2.8-4.4mmol/L, protein 0.15-0.45g/L, WCC (polymorph) to RCC ratio > 1:200, peripheral WCC > 11×10^9^/L).^19^ If the definition did not specify a cut-off for fever, we used a temperature > 38° C. Details regarding timing of EVD insertion and removal in relation to the timing of CSF sampling were applied when definitions specified these criteria. Variables that were subjective, such as headache, altered mental status, photophobia, nuchal rigidity and other clinical signs of meningitis that were not reliably obtainable in a critically ill population of patients due to mechanical ventilation, sedation or underlying disease process were excluded from 9 sets^20-28^ of diagnostic criteria. There were 3 sets of diagnostic criteria that contained data that are not routinely available, ^23, 29, 30^ the number of colony forming units/0.1 ml of CSF, the type of media the organism was grown on and antibody titres. The modified diagnostic criteria for VRI are shown in Supplementary Table 2.

### Statistical methods

Demographic data and clinical outcomes are presented as mean ± standard deviation for normally distributed continuous data, or median (interquartile range) for non-normally distributed data. Counts and proportions (of available data) was used for dichotomous data. Statistical analyses were conducted with chi-square tests for categorical variables, and one-way ANOVA or Kruskal-Wallis test for continuous variables. P values less than 0.05 were deemed to be significant. Bonferroni correction was applied where multiple testing occurred. All statistical analyses were performed using R (version 4.0.2, R Core Team, Vienna, Austria)

## Results

The flow of study participants is shown in Figure 1. There were 730 patients who were admitted with TBI, SAH and ICH to the neuroscience ICU between January 1^st^ 2015 to July 22^nd^ 2018. Of those patients, 10 did not consent to have their information used for research purposes. Of those remaining, there were 240 patients who had an EVD during admission and 190 with at least one CSF sample collected from the EVD.

**Figure 1.**
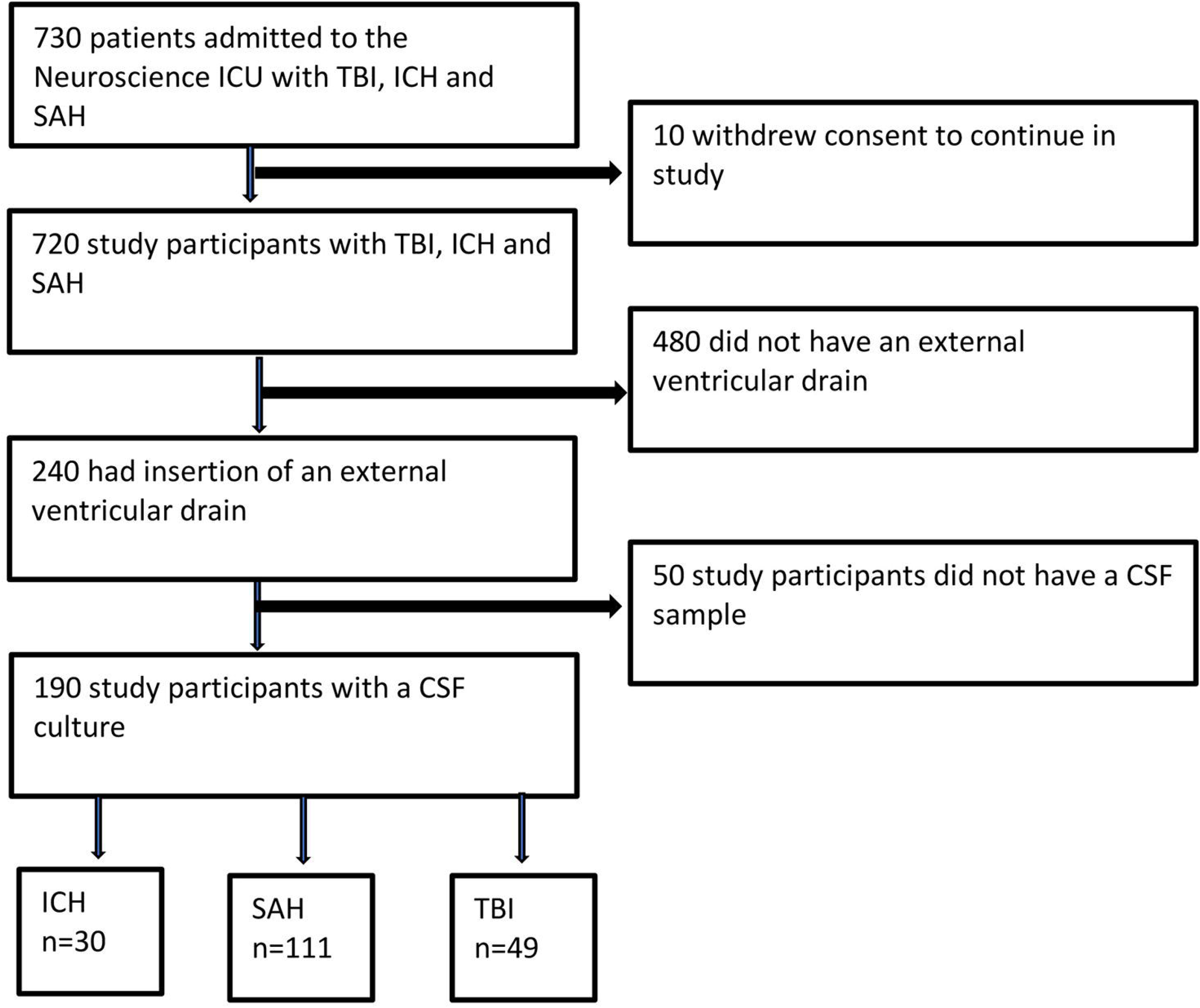
Flow of included patients ICU = intensive care unit, TBI = traumatic brain injury, SAH= subarachnoid haemorrhage, ICH = intracerebral haemorrhage, CSF = cerebrospinal fluid

The demographics and clinical characteristics of the included population are shown in Table 1. Invasive mechanical ventilation was required in 158 (83.2%) of our patient cohort, with a median (interquartile range (IQR)) duration of ventilation of 6 (IQR 2-13) days. The median (IQR) number of days receiving sedative agents was 3 (2 to 9) The median duration of EVD placement was 12 (IQR 7-17). Additional details regarding the severity of illness specific to the subgroups with ICH, SAH and TBI are shown in Supplemental Table 1.

**Table 1.**
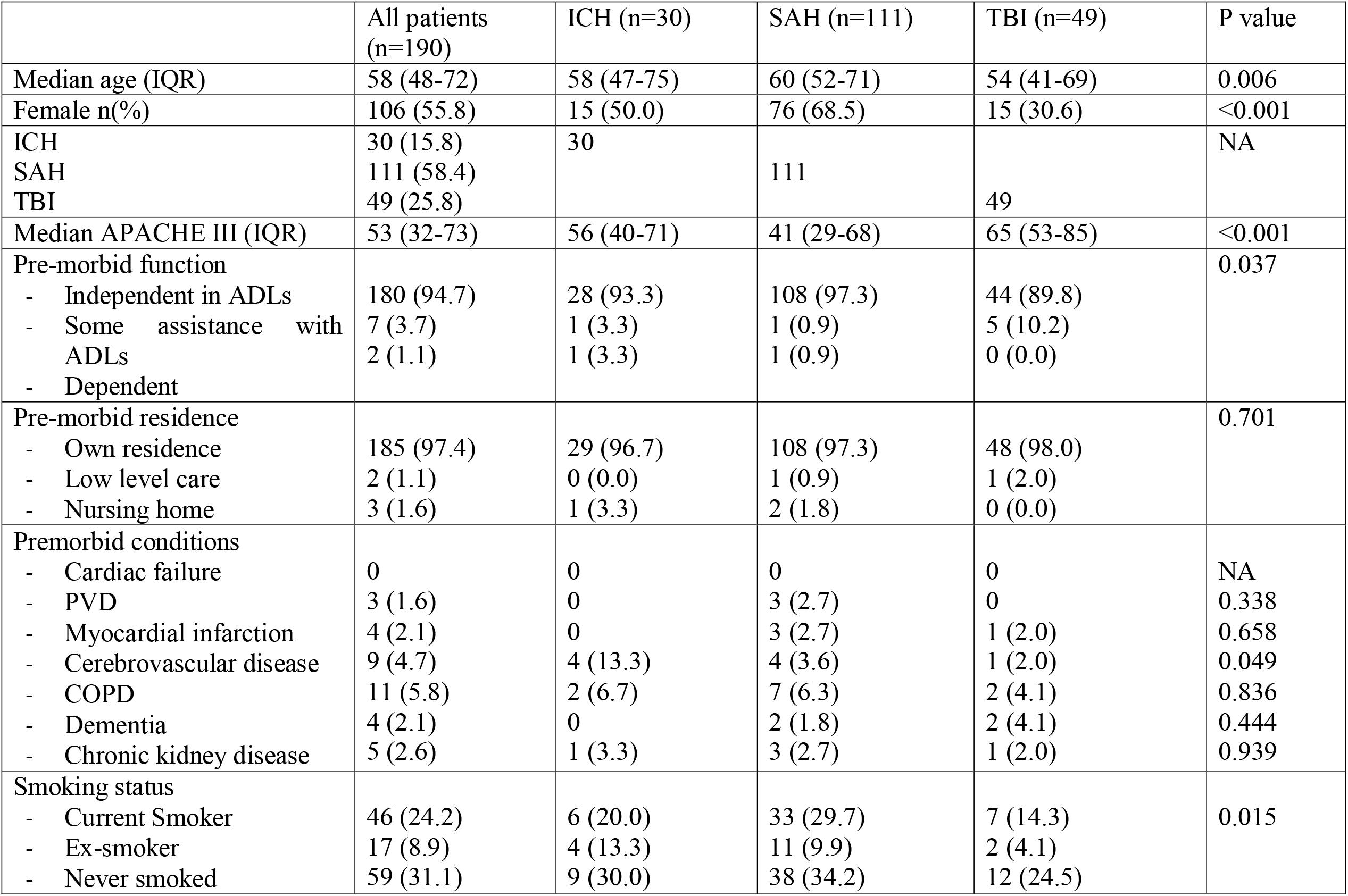
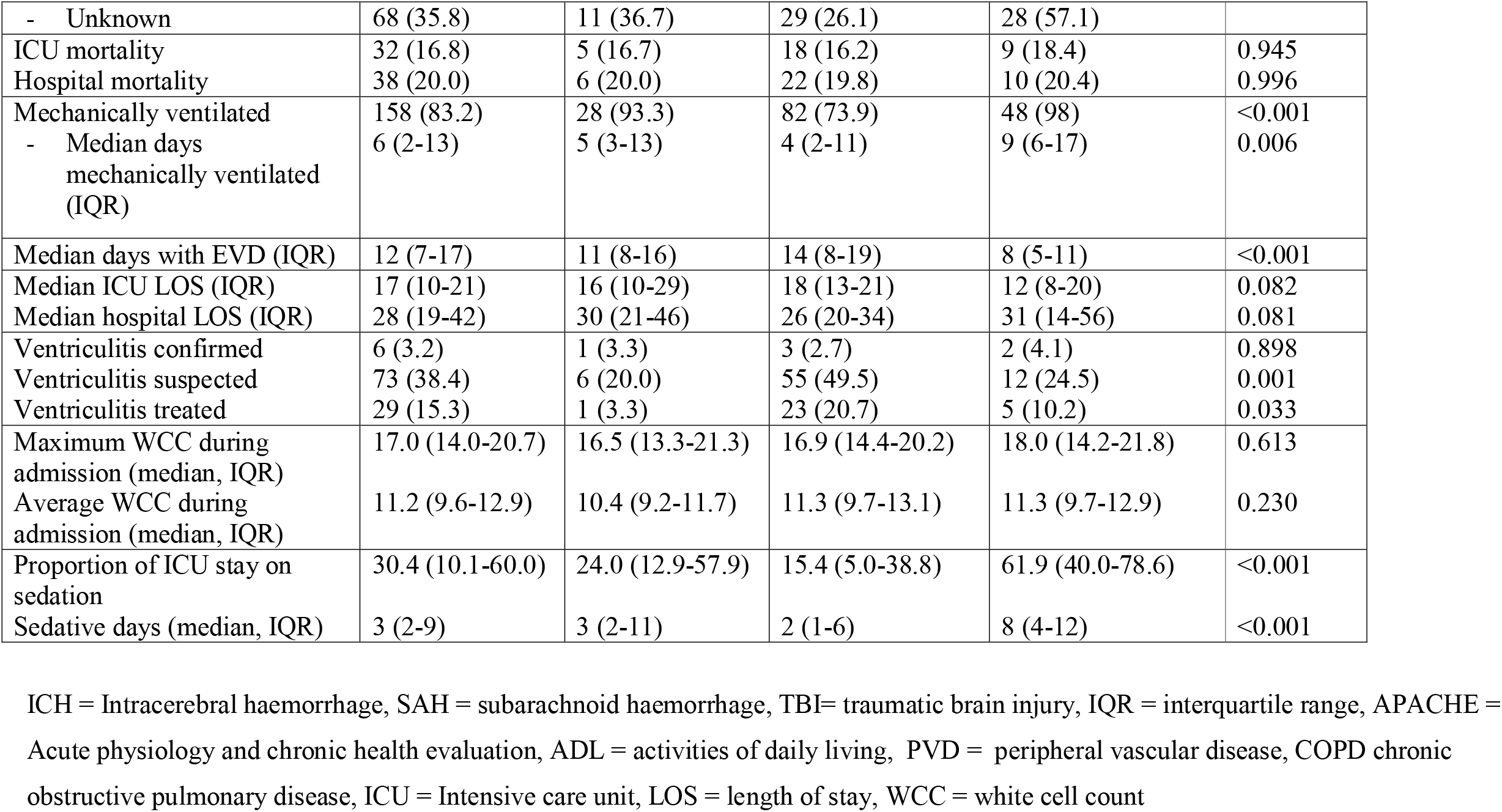
Characteristics of the cohort

The clinical cohort included 6 (3.2%) cases where a diagnosis of VRI was confirmed with a positive CSF culture. There were 29 (15.3%) participants who were treated for VRI. The clinical, laboratory and microbiological details of those with a confirmed diagnosis of VRI and those treated for VRI are shown in Table 2.

**Table 2.** Clinical, and laboratory features of clinical cases of VRI confirmed by positive culture, treated cases and suspected cases.

|  | Culture positive VRI (n=6) | Treated VRI (n=29) | Suspected VRI (n=73) | No suspicion of VRI (n=112) | P value |
| --- | --- | --- | --- | --- | --- |
| Age | 55 ± 17 | 55 ± 17 | 58 ± 14 | 58 ± 16 | 0.866 |
| Male | 4 (67) | 13 (45) | 25 (34) | 57 (51) | 0.106 |
| Primary pathology |  |  |  |  | 0.001 |
| ICH | 1 (17) | 1 (3) | 6 (8) | 24 (21) |  |
| SAH | 3 (50) | 23 (79) | 55 (75) | 52 (46) |  |
| TBI | 2 (33) | 5 (17) | 12 (16) | 36 (32) |  |
| Median APACHE III | 56 (40-68) | 37 (30-63) | 39 (29-63) | 61 (42-77) | 0.001 |
| Median EVD days | 18 (14-23) | 16 (13-23) | 17 (13-21) | 8 (4-13) | <0.001 |
| Ventilation on day of diagnosis n (%) | 3 (50) | 11 (38) | 34 (47) | 74 (67) | 0.010 |
| ICU LOS | 22 (19-33) | 20 (15-26) | 20 (15-28) | 13 (8-19) | <0.001 |
| Hospital LOS | 48 (32-60) | 30 (25-53) | 30 (24-47) | 24 (12-35) | <0.001 |
| Temperature (first day of diagnosis)* | 38.0 ± 0.8 | 38.3 ± 0.7 | 37.8 ± 0.7 | 37.0 ± 3.6 | <0.001 |
| WCC (first day of diagnosis)* | 8.0 (6.0-8.8) | 11.4 (9.1-15.0) | 12.9 (11.6-14.4) | 10.3 (8.4-14.1) | <0.001 |
| CRP first day of diagnosis* | 59 (38-198) | 31 (16-59) | 32 (17-90) | 12 (5-43) | 0.001 |
| CSF sample* |  |  |  |  |  |
| WCC | 330 (127-1424) | 141 (54-333) | 31 (10-138) | 22 (5-55) | <0.001 |
| RBC:WCC ratio | 52:1 | 63:1 | 60:1 | 257:1 | <0.001 |
| CSF glucose | 4.2 ± 0.8 | 4.0 ± 0.7 | 4.6 ± 0.6 | 4.8 ± 1.5 | 0.079 |
| Protein | 0.7 ± 0.4 | 0.7 ± 0.3 | 0.6 ± 0.4 | 1.4 ± 2.2 | 0.757 |
| Lactate | 3.6 ± 1.8 | 3.5 ± 0.8 | 2.9 ± 0.9 | 3.1 ± 1.6 | 0.216 |
| BSL* | 7.7 ± 1.4 | 7.4 ± 1.1 | 7.7 ± 1.3 | 8.2 ± 2.4 | 0.670 |
| Vasospasm | 1 (17) | 18 (62) | 35 (48) | 17 (15) | <0.001 |

VRI = ventriculostomy related infection, ICH = intracerebral haemorrhage, SAH = subarachnoid haemorrhage, TBI = traumatic brain injury, APACHE = Acute physiology and chronic health evaluation, EVD = external ventricular drain, ICU = intensive care unit, LOS = length of stay, WCC = white cell count, CRP = C reactive protein, CSF = cerebrospinal fluid, RBC = red blood cell, BSL = blood sugar level. \* For patients without a diagnosis or suspicion of VRI, the worst CSF sample results based on WCC and RBC:WCC ratio and the day this occurred were included for comparison

The estimate of the incidence of ventriculitis in the cohort varied from 0.5% to 95% when the various published sets of diagnostic criteria were used to define cases of VRI in this cohort, as shown in Table 3 and Figure 2.

**Table 3.**
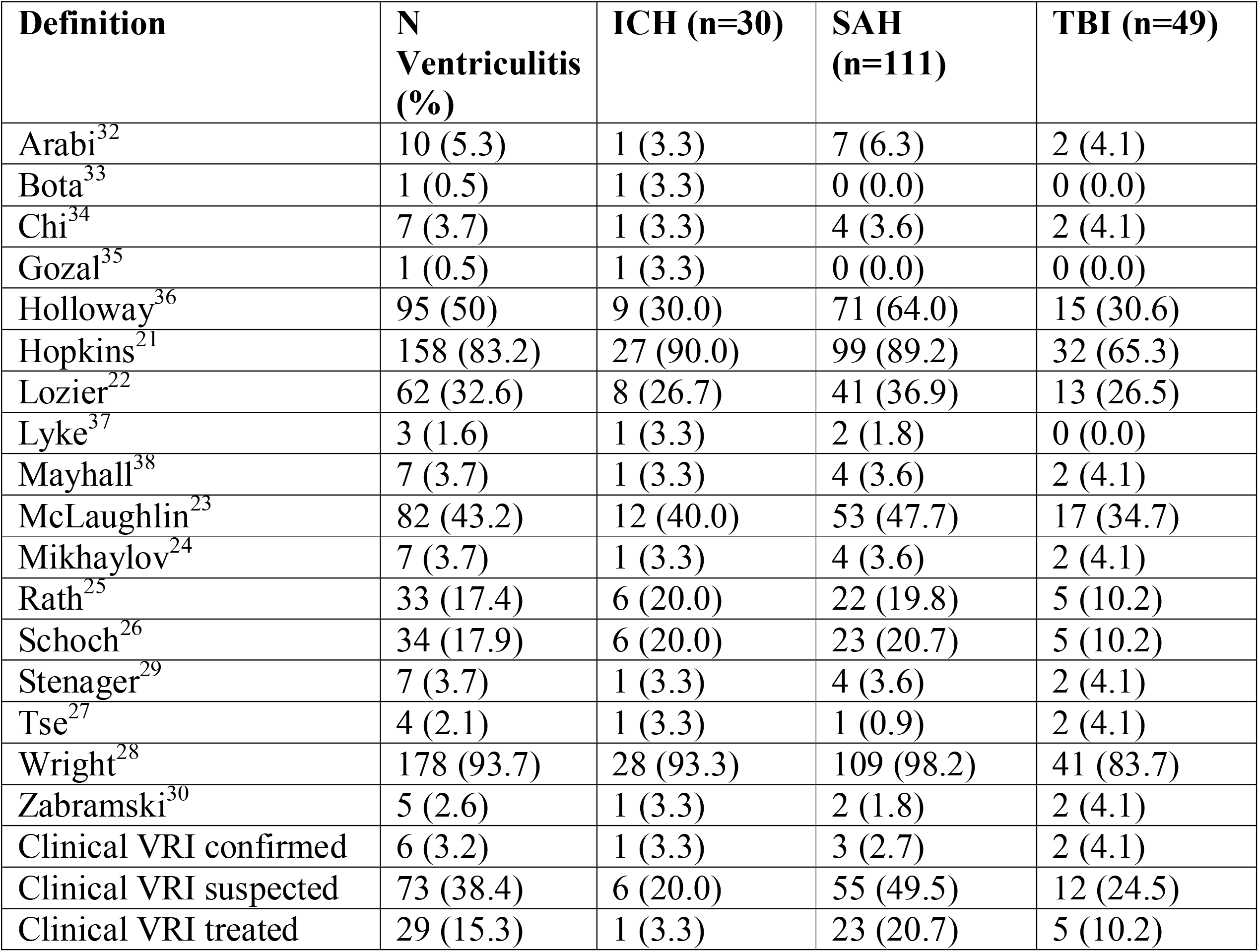
The incidence of ventriculitis according to the published definitions

| **Definition** | **N**<br>**Ventriculitis**<br>**(%)** | **ICH (n=30)** | **SAH**<br>**(n=111)** | **TBI (n=49)** |
| --- | --- | --- | --- | --- |
| Arabi<sup>32</sup> | 10 (5.3) | 1 (3.3) | 7 (6.3) | 2 (4.1) |
| Bota<sup>33</sup> | 1 (0.5) | 1 (3.3) | 0 (0.0) | 0 (0.0) |
| Chi<sup>34</sup> | 7 (3.7) | 1 (3.3) | 4 (3.6) | 2 (4.1) |
| Gozal<sup>35</sup> | 1 (0.5) | 1 (3.3) | 0 (0.0) | 0 (0.0) |
| Holloway<sup>36</sup> | 95 (50) | 9 (30.0) | 71 (64.0) | 15 (30.6) |
| Hopkins<sup>21</sup> | 158 (83.2) | 27 (90.0) | 99 (89.2) | 32 (65.3) |
| Lozier<sup>22</sup> | 62 (32.6) | 8 (26.7) | 41 (36.9) | 13 (26.5) |
| Lyke<sup>37</sup> | 3 (1.6) | 1 (3.3) | 2 (1.8) | 0 (0.0) |
| Mayhall<sup>38</sup> | 7 (3.7) | 1 (3.3) | 4 (3.6) | 2 (4.1) |
| McLaughlin<sup>23</sup> | 82 (43.2) | 12 (40.0) | 53 (47.7) | 17 (34.7) |
| Mikhaylov<sup>24</sup> | 7 (3.7) | 1 (3.3) | 4 (3.6) | 2 (4.1) |
| Rath<sup>25</sup> | 33 (17.4) | 6 (20.0) | 22 (19.8) | 5 (10.2) |
| Schoch<sup>26</sup> | 34 (17.9) | 6 (20.0) | 23 (20.7) | 5 (10.2) |
| Stenager<sup>29</sup> | 7 (3.7) | 1 (3.3) | 4 (3.6) | 2 (4.1) |
| Tse<sup>27</sup> | 4 (2.1) | 1 (3.3) | 1 (0.9) | 2 (4.1) |
| Wright<sup>28</sup> | 178 (93.7) | 28 (93.3) | 109 (98.2) | 41 (83.7) |
| Zabramski<sup>30</sup> | 5 (2.6) | 1 (3.3) | 2 (1.8) | 2 (4.1) |
| Clinical VRI confirmed | 6 (3.2) | 1 (3.3) | 3 (2.7) | 2 (4.1) |
| Clinical VRI suspected | 73 (38.4) | 6 (20.0) | 55 (49.5) | 12 (24.5) |
| Clinical VRI treated | 29 (15.3) | 1 (3.3) | 23 (20.7) | 5 (10.2) |

**Figure 2.**
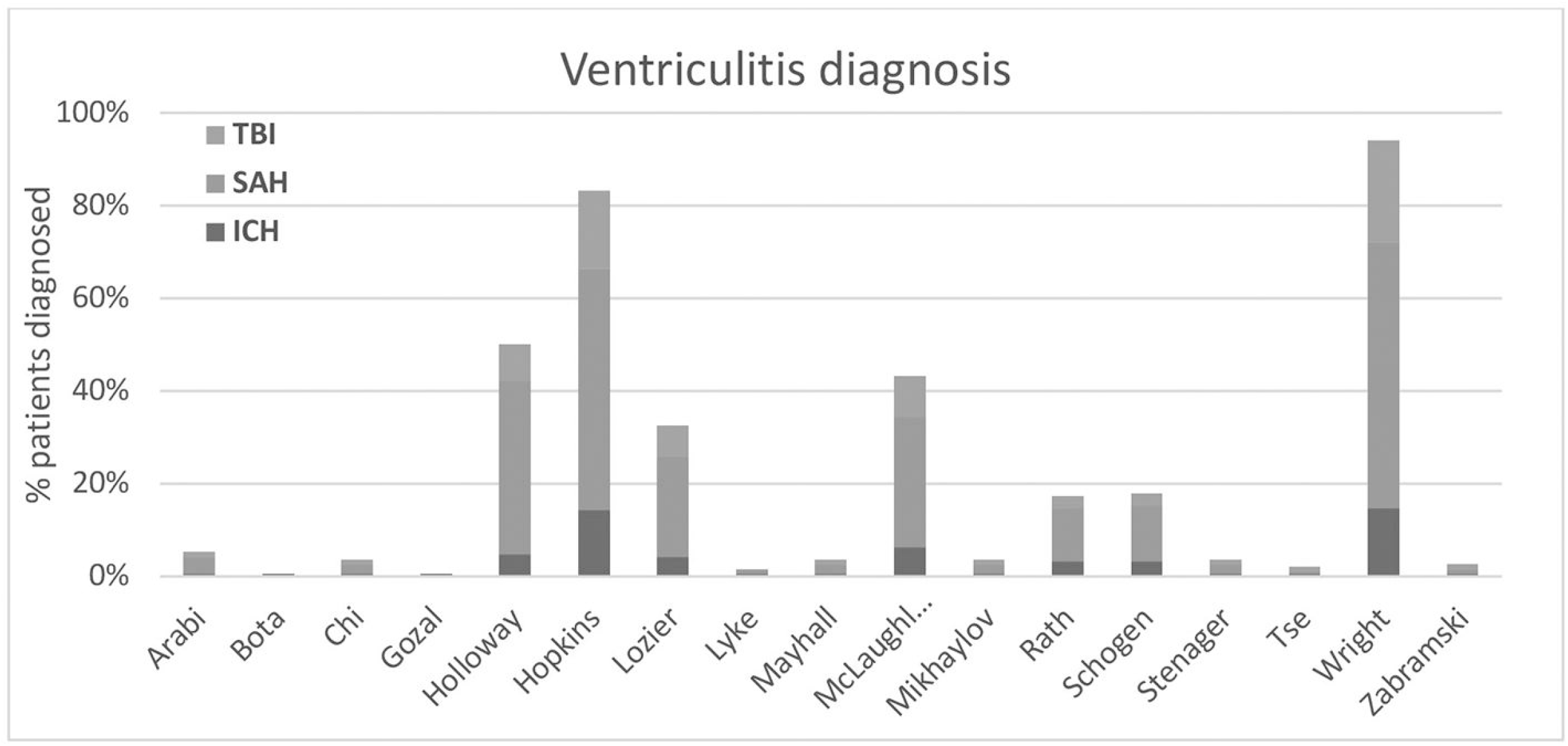
The incidence of ventriculitis in patients with ICH, TBI and SAH according to available published definitions.

## Discussion

We performed a retrospective cohort study to assess the variation in the incidence of VRI in a large cohort of patients with acute severe brain injury. We found that most patients with acute severe brain injury have a clinical condition or intervention such as a requirement for invasive mechanical ventilation that makes clinical parameters unreliable in the diagnostic assessment for VRI. We found an unacceptable degree of variation in the estimated incidence of VRI according to the published diagnostic criteria. The lack of clarity in defining a case of VRI was associated with a clinical impact, with a significant proportion of patients being treated with broad spectrum antimicrobial medications, in spite of failing to meet published diagnostic criteria for ventriculostomy related infection. A consistent, objective and universal set of diagnostic criteria for ventriculostomy related infection is needed.

In contrast to the previous published systematic review,^8^ our analysis also looked at the clinical impact of the variance in the definition by including patients treated for VRI, rather than simply including samples with a positive CSF culture. The results of our analysis are in accordance with the findings of Lewis et al, confirming wide variation in the estimated incidence of VRI.^8^ The proportion of patients treated for VRI in our cohort was in keeping with the findings of a previous systematic review that reported an incidence of VRI of approximately 23% across 42 studies assessing diagnostics criteria for VRI.^10^ In keeping with the results of that review, we excluded clinical factors that were known to be of limited diagnostic value in this critically ill population.^10^

There are a number of strengths to this study. We used prospectively collected data on consecutive, consenting participants to avoid selection bias. We used standard, a-priori definitions for cases of culture positive and treated cases of VRI, including only robust data points in these definitions. We included a diverse population of patients with SAH, TBI and ICH, representative of the population at risk for VRI in usual clinical practice. There are also a number of limitations to this analysis. Only small numbers of cases with positive cultures were identified, and as a single centre study, the results of this study may not be generalisable to other settings.

The major implication of this study for researchers and clinicians is that a standard definition of VRI is required. Clinical decisions regarding the use of broad-spectrum antimicrobial agents, the requirement for replacement of the EVD and delays in placement of a permanent VP shunt all hinge on having an accurate diagnosis of VRI. The variation in the estimated incidence leads to uncertainty for clinicians and patients and increases unnecessary practice variation. A standard definition of VRI is also essential for researchers to design and conduct the well-designed studies of diagnostic accuracy that are required to further knowledge in this area.^10^ The implementation of a bundle to prevent EVD infection has been suggested to be a quality indicator for Neurocritical care units.^31^ For this measure to be widely implemented, a standard definition of VRI is required in order for units to be able to compare their performance.

## Conclusion

The multiple available definitions of VRI are associated with an unacceptable broad estimated incidence of the incidence of this condition. Clinical conditions and interventions are common in this cohort of patients and limit the utility of including these variables as part of a robust diagnostic criteria. An objective, consistent and universal definition of VRI is needed.

## Data Availability

Data produced in this study are available upon reasonable request to teh authors

## Acknowledgements

The authors would like to acknowledge the study participants and their families for consenting to participate in this study.

## Authors contributions

Sara N. Bassin,, Simon Chadwick,, Oliver Flower, Pierre Janin, and Anthony Delaney conceived the study and wrote the initial study protocol, with critical revisions and input from Jonathan Parkinson and Archie Darbar

Sara N. Bassin, Emily Fitzgerald and Sajeev Mahendran collected data. David Tian performed the statistical analysis.

Sara N. Bassin MD and Anthony Delaney wrote the initial draft of the manuscript that was revised for important intellectual content by David Tian, Simon Chadwick, Sajeev Mahendran, Oliver Flower, Johnathan Parkinson, Archie Darbar and Pierre Janin.

All authors have read and approved the final version of the manuscript.

